# Elevated polyreactive immunoglobulin G in immune mediated liver injuries with the need for immunosuppressive therapy

**DOI:** 10.1101/2024.05.04.24306873

**Authors:** Theresa Kirchner, George N. Dalekos, Kalliopi Zachou, Mercedes Robles-Díaz, Raúl J. Andrade, Marcial Sebode, Ansgar Lohse, Maciej K. Janik, Piotr Milkiewicz, Mirjam Kolev, Nasser Semmo, Tony Bruns, Tom Jg Gevers, Benedetta Terziroli Beretta-Piccoli, Heiner Wedemeyer, Elmar Jaeckel, Richard Taubert, Bastian Engel, pIgG study group

## Abstract

**Background and aim:** The distinction of drug-induced liver injury (DILI), drug-induced autoimmune-like hepatitis (DI-ALH) and autoimmune hepatitis (AIH) can be challenging due to overlapping clinical characteristics. Recently, polyreactive immunoglobulin G (pIgG) was identified as a novel biomarker with a higher accuracy for the diagnose of AIH than conventional autoantibodies. This retrospective multicenter study aimed to evaluate the diagnostic accuracy of pIgG to distinguish between AIH, DI-ALH and DILI and thus identify patients in need of immunosuppression.

**Methods:** Samples from 116 patients (AIH=81, DI-ALH=12, DILI=23) were recruited and compared to a control group (non-AIH-non-DILI-LD= 596) from existing biorepositories.

**Results:** No patient in the DILI-group but 98% in the AIH-and 92% in the DI-ALH-group received immunosuppressive treatment. pIgG levels were significantly higher in the AIH-group (1.9 normalized arbitrary units (nAU) compared to DILI (1.1 nAU, p<0.001) and non-AIH-non-DILI-LD (1.0 nAU, p<0.001). Median pIgG concentrations of the DI-ALH-group (1.7 nAU) were between AIH (p=.634) and DILI (p=.052). Patients that needed immunosuppressive therapy for remission induction had significantly higher pIgG concentrations compared to those with spontaneous recovery of liver injury (1.8 nAU vs. 1.1 nAU, p<.001). The overall accuracy of pIgG >1.27nAU to distinguish AIH from DILI (74%) and liver injuries with and without the need for immunosuppression (74%) was similar to that of ANA (71/74%) and SMA (74/70%) at cut-offs of ≥1/40.

**Conclusion:** Polyreactive IgG can be used to predict AIH in comparison to DILI and indicate the need for immunosuppressive therapy in the work-up of immune mediated or drug-induced liver injuries.

## Introduction

Drug-induced liver injury (DILI) and autoimmune hepatitis (AIH) are difficult to differentiate from each other as they often share various clinical characteristics (1). AIH is a rare immune-mediated disease leading to cirrhosis and eventually liver-related death or liver transplantation when treated insufficiently, but when treated correctly prognosis is good in most of the cases (2). The diagnosis is based on autoantibody-measurements, histopathological findings and elevation of immunoglobulin G (IgG) (2). Continuous immunosuppressive therapy is needed to treat AIH and, in particular, to prevent the progression of liver disease (2). Although there is a diagnostic score (3) AIH remains a diagnosis of exclusion because there is no specific diagnostic marker.

Drug induced liver injury (DILI) can mimic the majority of other liver disorders and is triggered by various causative agents, for example antibiotics, metamizole, psychotropic drugs or herbal medication and dietary supplements (4, 5). Clinical presentation ranges from asymptomatic elevation of liver enzymes to acute liver failure (6). Idiosyncratic DILI affects only susceptible individuals and is less related to the drug dose (7). A temporal relationship with the suspected causative agent is needed and other competing etiologies have to be ruled out (5), however there is no specific diagnostic tool for DILI. Autoimmune features are rarely seen in DILI (8). After discontinuation of the causative agent DILI is a self-limiting disease with a good prognosis (5). Complementary to typical DILI, another drug related liver disease with autoimmune features is increasingly reported in the last years: drug-induced autoimmune like hepatitis (DI-ALH). DI-ALH is defined as a liver injury with laboratory and/or histological features that may be indistinguishable from idiopathic AIH (4). Previous studies showed that histological, biochemical and immunological features are overlapping in both immune-mediated entities (9).

Patients with AIH require long-term immunosuppressive therapy whereas immunosuppressive therapy can be safely withdrawn after weeks or a few months in DI-ALH (5). As distinction between AIH and DI-ALH is impossible in most cases at baseline based on the currently available diagnostic tools, immunosuppressive therapy is often started pragmatically and the final diagnosis can only be confirmed by the success or failure of an immunosuppression withdrawal attempt during follow-up (4).

Autoantibody measurement is a corner stone in the non-invasive diagnostic work-up of any unclear hepatitis. Conventional autoantibodies are antinuclear antibodies (ANA) and smooth-muscle antigen (SMA) antibodies in AIH type 1 while liver-kidney-microsomal (LKM) and/or liver-cytosolic type 1 antibodies are positive in AIH type 2. Soluble liver antigens (SLA) antibodies are positive in AIH type 3. However, there is a lack in sensitivity as well as in specificity for AIH causing diagnostic uncertainty (10,11,12).

Polyreactivity describes the potential of an autoantibody to bind multiple molecular structures, this leads to a higher affinity and neutralizing potential. Polyreactive immunoglobulins play an important role in primary immune response and in apoptosis. In 2022 polyreactive IgG (pIgG) was identified as a promising new biomarker to improve the diagnostic workup of any non-viral hepatitis with higher specificity and overall accuracy to distinguish AIH from non-AIH liver diseases than conventional autoantibodies in adults (13) and children (14) with additional value in autoantibody-negative AIH. However, DI-ALH and DILI were underrepresented in this initial characterization of pIgG (13).

This retrospective multicenter study aims to evaluate the diagnostic capacity of pIgG to predict AIH in comparison to DI-ALH, DILI and non-AIH-non-DILI liver diseases (non-AIH-non-DILI-LD). The distinction is clinically important to withhold immunosuppressive treatment in patients with DILI that do not need such treatment and differentiate AIH from DI-ALH to identify patients in whom immunosuppressive treatment may be safely discontinued.

## Patients and Methods

### Definitions

#### AIH

In accordance with clinical guidelines the simplified AIH score was ≥6 (3) in every case and the diagnosis was biopsy proven by compatible features. Clinical diagnosis and disease course were compatible with AIH and patients were dependent on immunosuppressive therapy longer than six months after diagnosis (15).

#### DI-ALH

DI-ALH was diagnosed in accordance with the expert opinion published by Andrade et al in 2023 (4). For DI-ALH there was a suspected causative agent and laboratory and histological features were comparable with the recently published expert opinion. Immunosuppressive therapy was initiated for remission induction in 92% of the cases, but there was no dependency on immunosuppressive medication for more than six months after the diagnosis (4). There was no relapse till the end of the study.

#### DILI

Definition was based on the latest EASL guideline for DILI (16). DILI was characterized by the existence of a suspected causative agent and the clinical and histological features were compatible with DILI. There was no need for immunosuppressive therapy at any time from diagnosis to last follow-up.

### Study population

116 adult patients (age ≥18 years) with AIH (n=81), DI-ALH (n=12) and DILI (n=23) without pre-existing liver disease from nine European centers were newly recruited for this retrospective multicenter study from existing biorepositories. Study groups were compared to an already established non-AIH-non-DILI-liver disease (non-AIH-non-DILI-LD) control group from a previous study (13). The non-AIH-non-DILI-LD group (n=596) particularly included alcoholic liver disease (n=90), primary sclerosing cholangitis (n=147), non-alcoholic fatty liver disease (n=204) and primary biliary cholangitis (n=125).

All included patients underwent a diagnostic liver biopsy for the work-up of unclear hepatitis. Serum samples were stored directly after onset and before the beginning of immunosuppressive treatment in every case. Samples were stored between 1990 and 2023. Patients with a loss to follow up within the first six months after liver biopsy or on immunosuppressive therapy prior to the liver biopsy were excluded.

### Quantification of polyreactive immunoglobulin G

Samples were stored at ≤ -20°C at Hanover Medical School. Samples from the other centers were cryo-conserved locally and sent frozen to Hanover. Quantification of pIgG was done using a custom-made Enzyme-linked Immunosorbent Assay (ELISA) with reactivity against human huntingtin-interacting protein 1-related protein (autoantigen) in bovine serum albumin blocked ELISA (HIP1R/BSA) in a single 1:100 dilution as published recently (13, 17). A standard curve was computed from five reference samples to calculate arbitrary units (AU) for each sample from the standard curve. AUs were normalized for center background and storage duration as published (normalized AU: nAU) (13). The test is outlined in more detail in the supplementary material. Autoantibody measurement was performed according to local standards in the participating centers in accordance with clinical guidelines (15, 16).

### Ethics

Written informed consent was obtained from all patients at respective centers. Use of material and data from all patients in this multicenter study was approved by the respective local ethical committees. The study was approved by the local Ethics Committee (Number 5582 with last Update from 2018, Hannover Medical School Ethics Committee, Hannover Medical School, Hannover, Germany). The study conforms to the ethical guidelines of the 1975 Declaration of Helsinki.

### Statistical analysis

Statistical analysis was performed using SPSS (version 15.0, SOSS; Inc, Chicago, IL), GraphPad Prism (version 10; GraphPad Prism Software Inc., La Jolla, CA) and Microsoft Excel (version 2019, Redmond, Washington).

Categorical variables are expressed as numbers and percentages; continuous variables are expressed as median and range. Chi²-test was used to compare contingency tables. The Mann-Whitney-U test was used to compare quantitative data between two groups and the Kruskal-Wallis-test was used to compare quantitative data between more than two groups. Area under the receiver operating characteristic (AUROC) analyses and Youden’s index were used to identify cut-off values. Accuracy of the diagnostic test was calculated as: (true positive + true negative)/total number. Sensitivities and specificities were compared with McNemar’s test.

P-values <0.05 (two-tailed) were considered significant in all analyses.

## Results

### Patient characteristics

This multicenter analysis included 81 AIH-, 12 DI-ALH-and 23 DILI-patients, that were newly recruited for this study, as well as a previously published comparator cohort of 596 non-AIH-non-DILI-LD patients taken from our previous study (13). Table 1 summarizes the main demographic and laboratory features of the study population. The median age was comparable between the groups (p=.164). The proportion of females was significantly higher in immune mediated liver diseases (AIH 79%, DI-ALH 75%) compared to DILI (57%) and non-AIH-non-DILI-LD (55%, p<0.001). AST and ALT were highest in DI-ALH and lowest in non-AIH-non-DILI-LD (both p<.001). IgG elevation in AIH (1.06 xULN) was higher compared to DI-ALH (0.89 xULN), DILI (0.75 xULN) and non-AIH-non-DILI-LD (0.80 xULN, p<.001, Table 1).

**Table 1:**
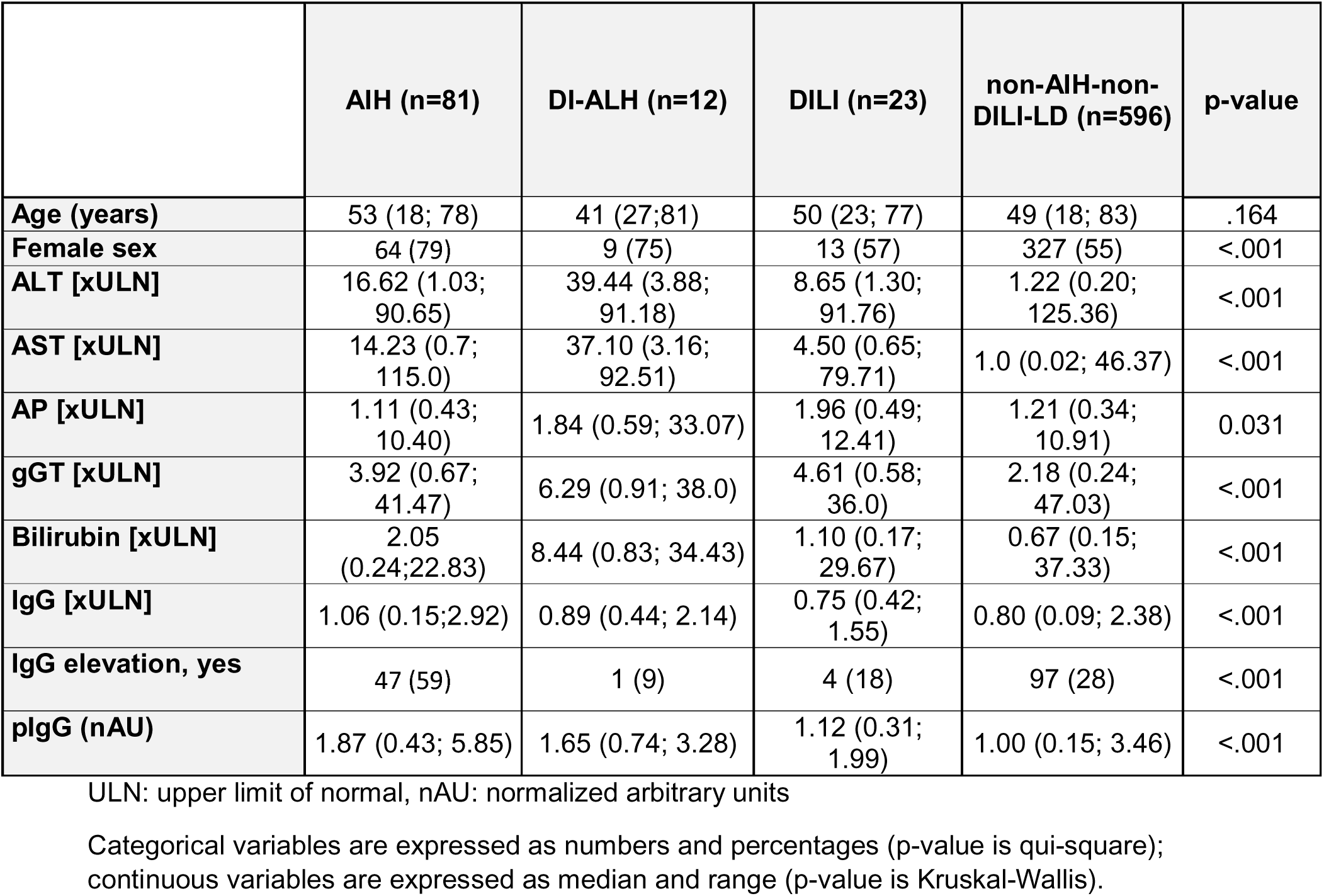
Baseline characteristics.

Suspected causative agents for DILI and DI-ALH are shown in Suppl. Table 1. Antibiotics and metamizole were the causative agent in most cases. Because of polypharmacy, the agent remained unknown in 17% of patients in both groups.

### Immunosuppressive treatment in AIH, DI-ALH and DILI

Ninety-eight percent of patients (n=79) in the AIH-group were treated with immunosuppressive therapy for remission induction (Suppl. Table 2). One patient refused to take the recommended medication and another had spontaneous remission without any therapy. One patient in the DI-ALH group refused to take the recommended immunosuppressive medication, therefore 92% of the DI-ALH group received immunosuppressive medication for remission induction. According to the given definitions no patient in the DILI-group received immunosuppressive therapy. In most cases corticosteroid monotherapy (51%) was initiated followed by a combination of steroids and mycophenolate mofetil in 28% in the AIH-group. At month six, 98% of patients in the AIH-group were treated with immunosuppressive therapy while therapy was discontinued in 100% of patients in the DI-ALH group. Immunosuppressive therapy changed from steroid monotherapy for remission induction to combination with MMF or azathioprine in most of the cases in the AIH-group. Comparative patient data are summarized in Suppl. Table 2.

### Polyreactive IgG was highest in AIH

The median pIgG level was highest in the AIH-group (1.87 nAU, range 0.43 - 5.85 nAU, Fig. 1A) and significantly higher compared to the DILI-(1.12 nAU, range 0.31 - 1.99 nAU) and the non-AIH-non-DILI-LD-group (1.00 nAU, range 0.15 - 3.46 nAU, both p<0.001). The median pIgG level in the DI-ALH (1.65 nAU, range 0.74-3.28 nAU) was significantly higher compared to the non-AIH-non-DILI-LD-group (p=.001). Statistical significance was narrowly missed for DI-ALH vs. DILI (p=0.052). Aside from that the difference between AIH and DI-ALH was not significant (p=.634, Fig. 1A).

**Figure 1.**
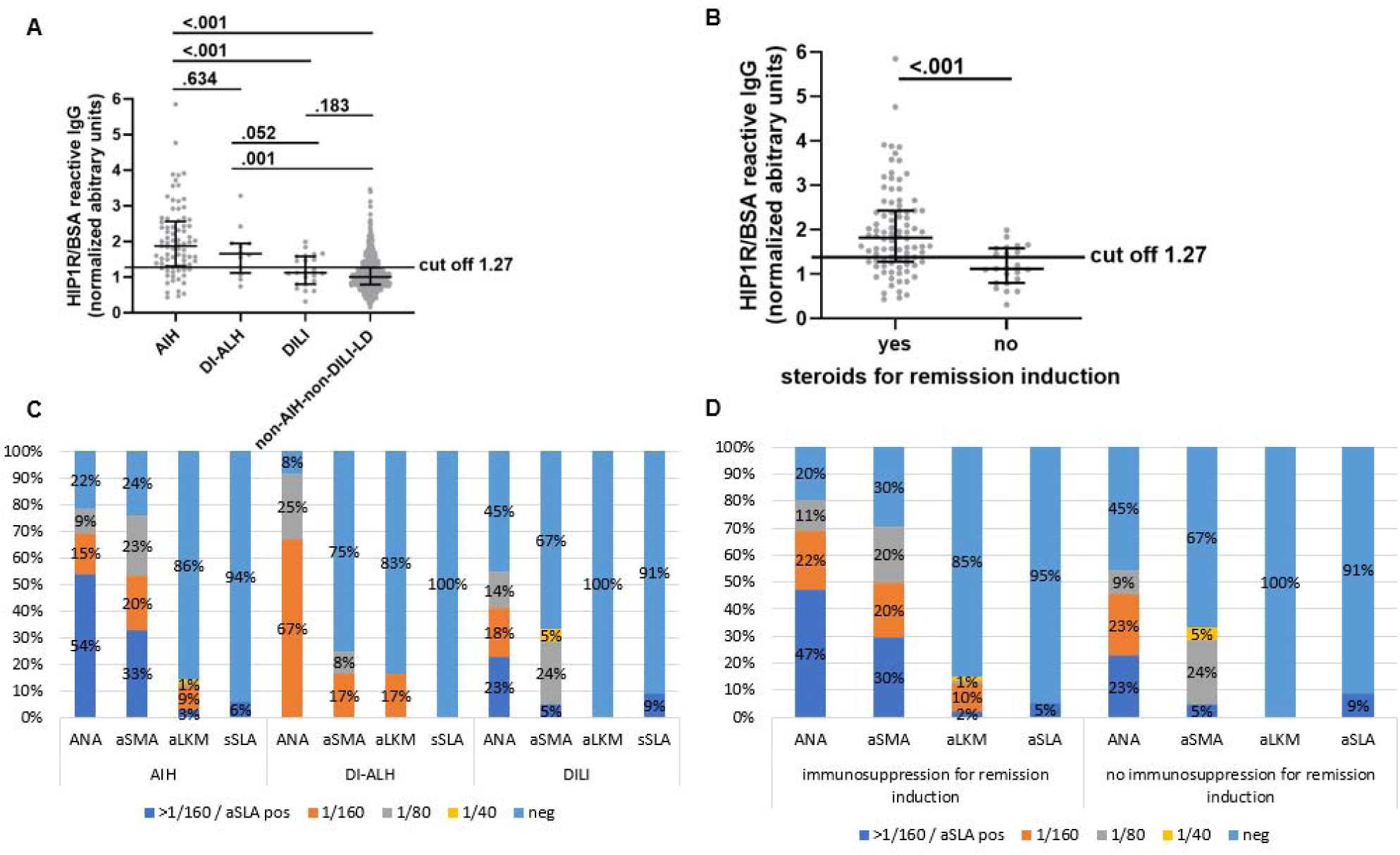
Overall diagnostic fidelity of HIP1R/BSA reactive IgG. Concentration of pIgG, expressed as nAU, in the different groups (AIH, DI-ALH, DILI and non-AIH-non-DILI-LD) (A) and in patients with and without immunosuppressive therapy for remission induction (B). The cut off of 1.27 was recently published for the distinction between untreated AIH and non-AIH diseases. Positivity and titer for conventional autoantibodies in the different groups (C) and in patients with and without immunosuppressive therapy for remission induction (D). Abbrevations: ANA antinuclear antibodies, aSMA anti smooth muscle antigen antibodies, aLKM anti liver kidney microsomal antibodies, aSLA anti soluble liver antigen antibodies.

When comparing all cases with immunosuppressive therapy for remission induction in the AIH-and the DI-ALH-group (n=90) to those liver injuries without immunosuppressive therapy for remission induction (DILI group, n=23) pIgG levels were significantly higher in patients with the need for immunosuppressive therapy (1.82 nAU (range 0.43-5.85 nAU) vs. 1.12 nAU (range 0.31 – 1.99 nAU), p<.001, Fig. 1B).

### Autoantibody measurement and serological features in AIH, DI-ALH and DILI

Positive serology (any positivity for ANA/aSMA/aLKM ≥1/40 and/or positivity for aSLA) was significantly more frequent in AIH (96%) and DI-ALH (92%) compared to DILI (70%, p=.001). The frequency of any antibody positivity was comparable between AIH and DI-ALH (p=.683). Positivity for ANA was highest in the DI-ALH group (92%) compared to the AIH-(78%) and the DILI-group (55%, p=.106, Fig. 1C). Titers for ANA were significantly higher in the AIH-and DI-ALH group (p<.001). Staining pattern was fine speckled (AIH and DI-ALH both 46%), homogenous (AIH 23%, DI-ALH 9%) and nucleolar (AIH 5%, DI-ALH 37%) in most of the immune-mediated cases. Highest positivity for aSMA was seen in the AIH-group (76%, p<.001). In addition, there were higher aSMA titers in AIH compared to the other groups (median AIH ≥1/160 (range 1/80 - ≥1/160) vs DI-ALH 1/160 (range 1/80 - 1/160) vs DILI 1/80 (range 1/40 - ≥1/160), p<.001). Positivity for aSMA in the DI-ALH-and DILI-group was comparable (25% vs 33%, p=0.635). Positivity for aLKM was comparable in the AIH-(14%) and DI-ALH-group (17%) whereas all patients in the DILI-group where negative for aLKM. aSLA was negative in every DI-ALH case and showed low and comparable positivity in the AIH-(6%) and the DILI-group (9%, Fig. 1C).

When comparing patients with immunosuppression for remission induction and those without immunosuppressive therapy a positive autoantibody serology (any ANA, SMA, LKM or SLA, according to the given definition) could be seen in 97% of the patients with therapy and in 70% in the cases without therapy (p<.001). Positivity for ANA was 80% in the therapy-group compared to 55% in the group without therapy (p=.012, Fig 1D). ANA-titers were comparable in both groups (median immunosuppressive treatment ≥1/160 (median 1/80 - ≥1/160 vs no immunosuppressive therapy 1/160 (range 1/80 - ≥1/160, p=.088). Positivity for aSMA was significantly higher in the treatment group (70% vs 33%, p=.002). Median aSMA titer was ≥1/160 (range 1/80 - ≥1/160) in the treatment and 1/80 (range 1/40 - ≥1/160) in the no treatment group. aLKM was negative in every case without immunosuppression and positivity for aSLA was comparable (5% vs 9%, p=.628).

### Overall diagnostic fidelity of pIgG and the autoantibodies

As published recently (13) a cut off of 1.27 nAU for positivity of pIgG and a cut off ≥1/40 for positivity in autoantibody measurement were used to evaluate the diagnostic fidelity of pIgG in comparison to conventional autoantibody measurement in this study. When comparing AIH to DILI with these cut-offs, sensitivity of pIgG (79%), ANA (78%, p=.652) and aSMA (76%, p=.521) were comparable, but significantly lower for aLKM (14%, p=.002) and aSLA (6%, p=.012, Fig. 2A and Suppl. Table. 3). Specificity of aLKM (100%, p=.002) and aSLA (91%,p=.015) were higher compared to pIgG (61%) and aSMA (67%, p=.549). The lowest specificity was seen for ANA (46%, p= .042). Overall accuracy was highest for pIgG and aSMA (both 74%) and comparable to ANA (71%). Overall accuracy was 32% for LKM and 18% for aSLA.

**Figure 2.**
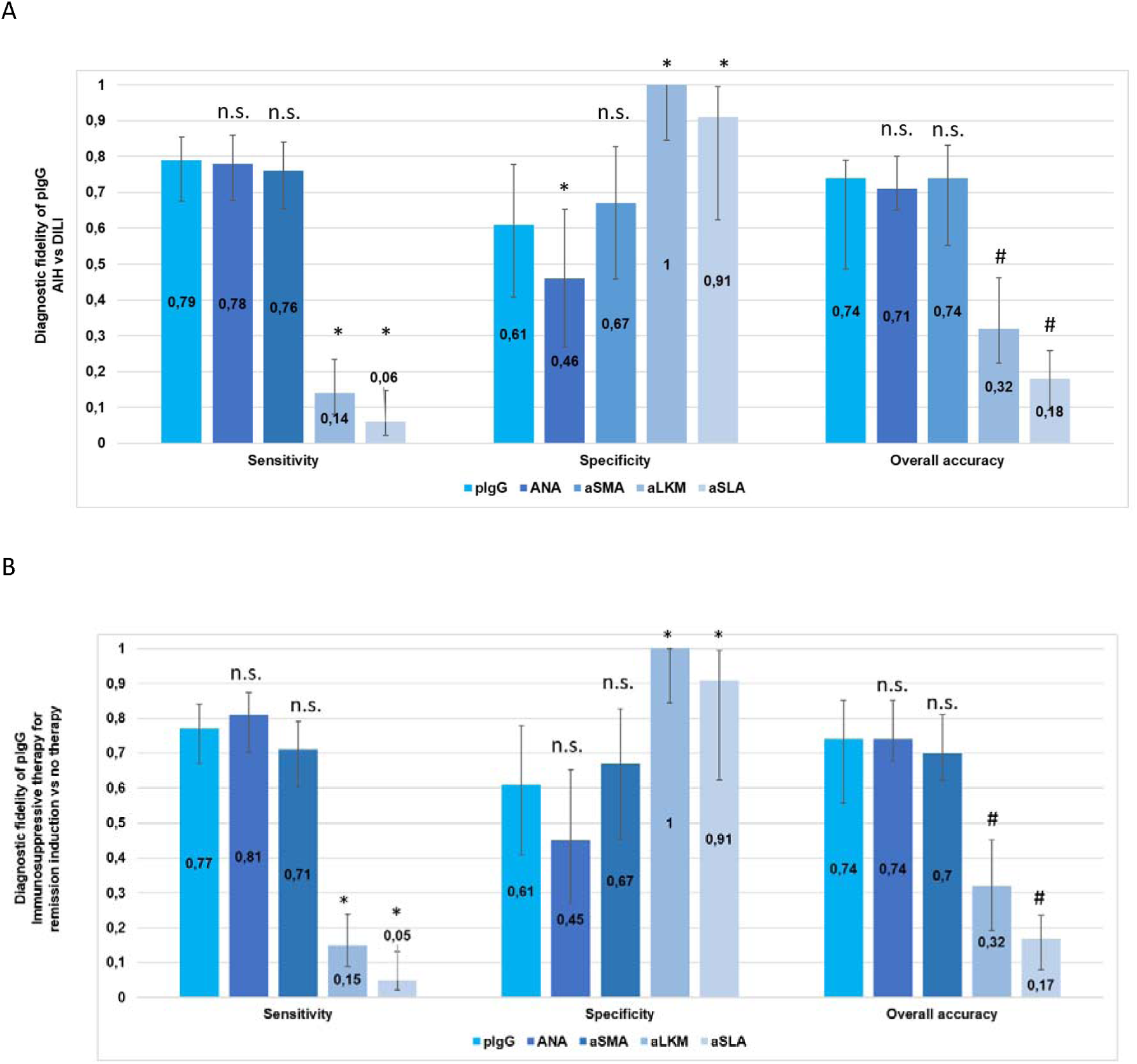
Overall diagnostic fidelity of pIgG and conventional autoantibodies to distinguish untreated AIH from DILI (A) and to distinguish patients with immunosuppressive therapy for remission induction from those without therapy (B). Error bars indicate 95% CI. * p<0.05 for McNemaŕs test comparing pIgG (cut off 1.27nAU) to other autoantibodies, # significant difference to pIgG by missing overlap of the 95% CI when McNemaŕs test was not applicable, n.s.: non significant, p≥ 0.05.

We hypothesized that the identification of new and higher cut-off might increase the accuracy for the distinction of AIH and DILI similar to different titers of conventional autoantibodies. Using Youden’s index, a new cut off (1.71 nAU) for positivity of pIgG was evaluated in this cohort to improve the discrimination of AIH from DILI. Sensitivity was 56% and specificity was 91%, while overall accuracy was 64%. In comparison, the modified new cut off for this cohort reached a higher specificity with lower overall accuracy (Suppl. Table 3). Likewise, a new cut off of ≥1/160 for ANA and 1/160 for SMA was determined using Youden’s index to improve specificity. Using this new cut off sensitivity of ANA (69%) and aSMA (70%) declined, while specificity was higher (ANA 58%, aSMA 86%). Comparable to pIgG, the overall accuracy for ANA (67%) and aSMA (71%) was lower using modified cut offs. In summary, no higher cut-off improved the overall accuracy of any autoantibody including pIgG in this study.

When comparing the diagnostic fidelity of pIgG and the conventional autoantibodies to distinguish between patients that needed immunosuppression for remission induction and patients without immunosuppressive therapy, sensitivity of pIgG (cut off of 1.27 nAU, 77%) was not significantly different to ANA (81%, p=.721) and aSMA (71%, p=.830, Fig. 2B and Suppl. Table 3). Sensitivity was lowest for aLKM (15%, p=.012) and aSLA (5%, p=.005). Specificity was comparable for pIgG (61%) and aSMA (67%, p=.589) and lower for ANA (45%, p=.720). Highest specificity was reached for aLKM (100%, p=.037) and aSLA (91%, p=.014). Overall accuracy for pIgG (74%), ANA (74%) and aSMA (70%) were comparable and significantly lower for aLKM (32%) and aSLA (17%).

We tried to identify a new and higher cut-off to improve the discrimination of patients with liver injury with and without the need for immunosuppression. A higher cut-off (2.00 nAU) increased the specificity (100%), but decreased the sensitivity (40%) and overall accuracy (66%, Suppl. Fig. 3B). A higher cut off for ANA with ≥1/160 and aSMA with 1/160 leads to a higher specificity and lower sensitivity and overall accuracy, too.

## Discussion

Diagnosing unclear hepatitis can be challenging because immune-mediated liver diseases with a need for immunosuppressive therapy for remission induction and self-limiting liver diseases often share similar clinical characteristics. Testing of autoantibodies using the current gold standard of immunofluorescence (IFT, 18) and liver histopathology is time consuming. There is a need for a non-invasive, cheap and quickly available biomarker to distinguish these entities and to support decision making for or against immunosuppressive therapy.

In 2022, we proposed pIgG as a new diagnostic biomarker for AIH demonstrating higher specificity and greater overall accuracy compared to conventional autoantibodies (13). Reactivity in a HIP1R/BSA ELISA was outlined as a surrogate marker for pIgG concentration. In this study, DILI were underrepresented and DI-ALH was not yet defined, so AIH and DI-ALH were summarized under the label AIH and a retrospective distinction was not possible. As a consequence, the aim of the present study was to include more clearly defined cases of DILI and DI-ALH in addition to a cohort of AIH and non-AIH-non-DILI liver diseases.

In summary, pIgG was significantly elevated in AIH compared to DILI while DI-ALH had pIgG levels between both groups. In this cohort significance was missed in the comparison of DI-ALH and DILI (p=0.052), most likely owing due to small sample number which is a limitation of the current study. However, at baseline, during the work-up of an unclear hepatitis, it is more important to get guidance regarding the need to start an immunosuppression to induce remission rather than having the exact discrimination between AIH and DI-ALH. The finding, that pIgG levels are significantly increased in those liver injuries, in which the treating physicians saw a need to start immunosuppression, is even more important. The obvious limitation of this parameter is that it was not made within a standardized study concept or by a central board. However, all the contributing centers were high volume expert centers mostly coming from the European Reference Network for rare liver diseases (ERN Rare Liver).

The initial analyses were made with the pIgG cut-off for the distinction between AIH and non-AIH-liver diseases from our previous study (1.27 nAU). As for many diagnostic tests, the specificity of the conventional autoantibodies for the diagnosis of AIH could be improved by higher thresholds of the antibody titers as shown in detail most recently (19). So, we also tried to identify a higher and more specific pIgG cut-off for the distinction of AIH and DILI and for liver injuries with/without the need for immunosuppression. This modified higher cut-off for the positivity of pIgG lead to a higher specificity but at the cost of a lower sensitivity and overall accuracy. The same improvement of specificity with less sensitivity and less overall accuracy could be observed for the conventional autoantibodies as well (19).

While pIgG had a superior accuracy for the distinction between AIH and many different non-AIH-liver diseases in adults and children (13, 14), pIgG exhibited comparable accuracies like ANA and aSMA but still higher accuracies than aLKM and aSLA (Fig. 1) for the concrete distinction between AIH and DILI or between liver injuries with and without the need for immunosuppression. Interestingly, the pIgG assay exhibited a similar accuracy in the present study (AIH vs DILI 74%; with/without need for immunosuppression 74%) as in the previous study (AIH vs. non-AIH liver diseases 73%) (13). However, ANA and aSMA exhibited a better overall performance in the current study (AIH vs DILI 71% and 74%; with/without need for immunosuppression 73% and 70%) compared to the previous study (AIH vs. non-AIH liver diseases 65% and 64%). Therefore the non-inferiority is not caused by a worse performance of the pIgG but by a better performance of the IFT in the present study.

Current markers are already good in the discrimination between immune-mediated liver diseases and DILI. Compared to the current gold standard of conventional autoantibody testing via IFT on rodent tissue sections or HEp2cells (15, 17, 18), pIgG are quantified via a solid phase assay with recombinant peptide being less labor-intensive. Aside from that the assay can be automated easily and is more objectively than IFT which is dependent on the examiner. Additionally, there is only one dilution step (1/100) in the pIgG assay and not a titration (1/40, 1/80, 1/160 etc.) which leads to less serum demand.

Limitations of this retrospective study are small sample numbers for DILI and DI-ALH due to the analysis of rare diseases and available biomaterial was even rarer. Additionally, important quality measures were implemented such as a mandatory liver biopsy and clinical follow-up regarding the short-and long-term dependency on immunosuppression in case of DILI and DI-ALH. The limitations of the start of the immunosuppression according to the local physicians was already discussed above. The current cohort also has the bias of non-standardized autoantibody measurement. We have just initiated a prospective multicenter study to validate the accuracy of pIgG for the prediction of AIH, DI-ALH, DILI and other non-viral liver diseases as well as the need for immunosuppression in liver injuries (NCT05810480). We expect the first results in two to three years from now.

In conclusion pIgG could be used as a promising additional biomarker to distinguish between AIH and DILI and for the prediction of an immunosuppression-dependent liver injury with similar accuracies like the conventional autoantibodies ANA and aSMA in IFT but with a less labor intense ELISA. A prospective evaluation of pIgG has been initiated in Europe, but the results are still pending.

## Abbreviations

AIH: Autoimmune hepatitis
AMA: anti-mitochondrial antibody
ANA: antinuclear antibody
DI-ALH: Drug-induced autoimmune-like hepatitis
DILI: Drug-induced liver injury
LKM: Liver kidney microsomal antibodies
Non-AIH-non-DILI-LD: non-autoimmune hepatitis non-drug-induced liver injury - liver disease
pIgG: polyreactive immunoglobulin G
SMA: smooth-muscle antigen
SLA: soluble liver antigens

## Ethics approval statement

Written informed consent was obtained from all patients at respective centers. Use of material and data from all patients in this multicenter study was approved by the respective local ethical committees. The study conforms to the ethical guidelines of the 1975 Declaration of Helsinki.

## Acknowledgements

We thank Stephanie Loges and Nicole Henjes from the Gastroenterology autoantibody laboratory at Hannover Medical School for their constant support during this project. We thank Konstantinos Iordanidis at Hannover Medical School for technical assistance in performing the experiments. We thank the International AIH Group (IAIHG) and the ERN Rare Liver for creating a platform for international research networking and collaboration.

## Writing Assistance

None

## Author contributions to manuscript

Study concept and design: Richard Taubert, Bastian Engel.

Acquisition of data: Theresa Kirchner, Richard Taubert, Bastian Engel, George N. Dalekos, Kalliopi Zachou, Mercedes Robles-Díaz, Raúl J. Andrade, Marcial Sebode, Ansgar Lohse, Maciej K. Janik, Piotr Milkiewicz, Mirjam Kolev, Tony Bruns, Nasser Semmo, Sarah Habes, Ye H Oo, Claudine Lalanne, Simon Pape, Joost P H Drenth, Luigi Muratori, Jessica K Dyson, Isabel Graupera, Benedetta Terziroli Beretta-Piccoli.

Drafting of the manuscript: Theresa Kirchner, Richard Taubert, Bastian Engel.

Critical revision of the manuscript for important intellectual content: George N. Dalekos, Kalliopi Zachou, Mercedes Robles-Díaz, Raúl J. Andrade, Marcial Sebode, Ansgar Lohse, Maciej K. Janik, Piotr Milkiewicz, Mirjam Kolev, Tony Bruns, Tom Jg Gevers, María-Carlota Londoño, Sarah Habes, Ye H Oo, Claudine Lalanne, Simon Pape, Joost P H Drenth, Luigi Muratori, Jessica K Dyson, Isabel Graupera,Benedetta Terzirlo Beretta-Piccoli, Heiner Wedemeyer, Elmar Jaeckel.

Statistical analysis: Theresa Kirchner, Richard Taubert, Bastian Engel.

Obtained funding: Richard Taubert, Bastian Engel.

Administrative, technical, or material support: Richard Taubert, Bastian Engel, Heiner Wedemeyer, Elmar Jaeckel.

Study supervision: Richard Taubert, Bastian Engel.

## Data availability statement

The data that support the plots within this paper and other findings of this study are available from the corresponding authors upon reasonable request.

---

**Suppl. Table 1:**
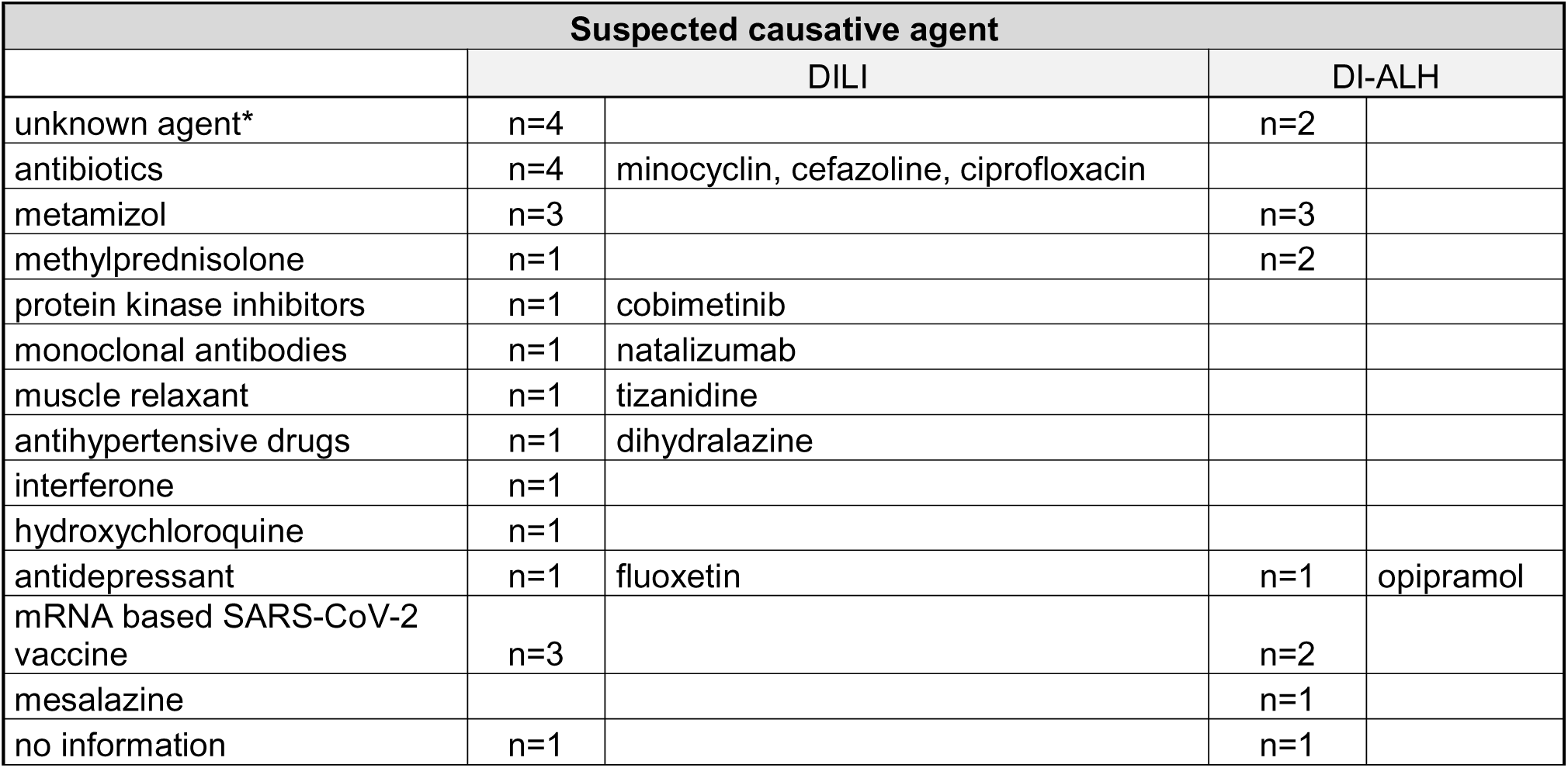
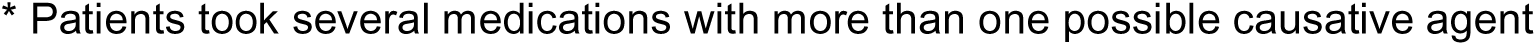
Suspected causative agents in DILI and DI-ALH.

| Suspected causative agent |  |  |  |  |
| --- | --- | --- | --- | --- |
|  | DILI |  | DI-ALH |  |
| unknown agent* | n=4 |  | n=2 |  |
| antibiotics | n=4 | minocyclin, cefazoline, ciprofloxacin |  |  |
| metamizol | n=3 |  | n=3 |  |
| methylprednisolone | n=1 |  | n=2 |  |
| protein kinase inhibitors | n=1 | cobimetinib |  |  |
| monoclonal antibodies | n=1 | natalizumab |  |  |
| muscle relaxant | n=1 | tizanidine |  |  |
| antihypertensive drugs | n=1 | dihydralazine |  |  |
| interferone | n=1 |  |  |  |
| hydroxychloroquine | n=1 |  |  |  |
| antidepressant | n=1 | fluoxetine | n=1 | opipramol |
| mRNA based SARS-CoV-2 vaccine | n=3 |  | n=2 |  |
| mesalazine |  |  | n=1 |  |
| no information | n=1 |  | n=1 |  |

\* Patients took several medications with more than one possible causative agent

**Suppl. Table 2:** Immunosuppressive therapy.

|  |  | **AIH**<br>**(n=81)** | **DI-ALH**<br>**(n=12)** | **DILI**<br>**(n=23)** | **p** |
| --- | --- | --- | --- | --- | --- |
|  |  | median<br>(min; max)<br>/ n (%) | median<br>(min; max)<br>/ n (%) | median<br>(min;<br>max) / n<br>(%) |  |
| **Simplified AIH Score\*** |  | 7 (6; 9) | 5 (4; 5) | 5 (2; 7) | <.001 |
| **Immunosuppressive**<br>**treatment for remission**<br>**induction** | **yes** | 79 (98) | 11 (92) | 0 (0) | <.001 |
| **Immunosuppressive**<br>**drug for remission**<br>**induction** | **steroids** | 40 (51) | 10 (100) |  | .067 |
|  | **steroids+MMF** | 22 (28) | 0 (0) |  |  |
|  | **steroids+azathioprine** | 9 (11) | 0 (0) |  |  |
|  | **steroids+azathioprine+MMF** | 1 (1) | 0 (0) |  |  |
|  | **unknown** | 7 (9) | 0 (0) |  |  |
| **Immunosuppressive**<br>**treatment at month 6** | **yes** | 79 (98) | 0 (0) | 0 (0) | <.001 |
| **Immunosuppressive**<br>**drug at month 6** | **steroids** | 27 (34) |  |  |  |
|  | **MMF** | 4 (5) |  |  |  |
|  | **steroids+MMF** | 17 (22) |  |  |  |
|  | **steroids+azathioprine** | 18 (23) |  |  |  |
|  | **other** | 3 (4) |  |  |  |
|  | **unknown** | 10 (13) |  |  |  |

\* Hennes et al, Simplified criteria for the diagnosis of autoimmune hepatitis. Hepatology. 2008 Combination of medication were prescribed sequentially.

**Suppl. Table 3:** Diagnostic fidelity for AIH vs DILI (3A) and immunosuppressive therapy vs no immunosuppressive therapy (3B) for pIgG, conventional autoantibodies and modified cut offs.

|  | Sensitivity | 95% CI | Specificity | 95% CI | Overall accuracy | 95% CI |
| --- | --- | --- | --- | --- | --- | --- |
| pIgG cut off 1.27 nAU | 0.778 | 0.676-0.855 | 0.609 | 0.408-0.778 | 0.740 | 0.486-0.791 |
| pIgG cut off 1.71 nAU | 0.556 | 0.447-0.659 | 0.913 | 0.732-0.985 | 0.635 | 0.525-0.789 |
| ANA cut off $\geq 1/40$ | 0.782 | 0.678-0.859 | 0.455 | 0.269-0.653 | 0.710 | 0.651-0.801 |
| ANA cut off $\geq 1/160$ | 0.689 | 0.564-0.790 | 0.583 | 0.320-0.807 | 0.671 | 0.492-0.718 |
| aSMA cut off $\geq 1/40$ | 0.760 | 0.655-0.840 | 0.667 | 0.458-0.828 | 0.740 | 0.552-0.832 |
| aSMA cut off 1/160 | 0.695 | 0.569-0.800 | 0.857 | 0.497-0.993 | 0.712 | 0.660-0.834 |
| aLKM cut off $\geq 1/40$ | 0.141 | 0.081-0.235 | 1.000 | 0.845-1.000 | 0.323 | 0.224-0.462 |
| aSLA | 0.060 | 0.023-0.148 | 0.909 | 0.623-0.995 | 0.180 | 0.092-0.259 |

**3B**
|  | Sensitivity | 95% CI | Specificity | 95% CI | Overall accuracy | 95% CI |
| --- | --- | --- | --- | --- | --- | --- |
| pIgG cut off 1.27 nAU | 0.767 | 0.670-0.842 | 0.609 | 0.408-0.778 | 0.735 | 0.557-0.852 |
| pIgG cut off 2.00 nAU | 0.400 | 0.305-0.503 | 1 | 0.857-1 | 0.656 | 0.437-0.802 |
| ANA cut off $\geq 1/40$ | 0.805 | 0.702-0.874 | 0.454 | 0.270-0.653 | 0.734 | 0.678-0.851 |
| ANA cut off $\geq 1/160$ | 0.769 | 0.698-0.912 | 0.545 | 0.374-0.687 | 0.661 | 0.597-0.781 |
| aSMA cut off $\geq 1/40$ | 0.705 | 0.602-0.790 | 0.667 | 0.454-0.828 | 0.700 | 0.621-0.812 |
| aSMA cut off 1/160 | 0.564 | 0.427-0.665 | 0.952 | 0.731-1 | 0.593 | 0.461-0.784 |
| aLKM cut off $\geq 1/40$ | 0.150 | 0.090-0.240 | 1.000 | 0.845-1.00 | 0.315 | 0.192-0.452 |
| aSLA | 0.054 | 0.021-0.131 | 0.909 | 0.623-0.995 | 0.165 | 0.078-0.235 |

