## Supplemental file for "Elevated polyreactive immunoglobulin G in immune mediated liver injuries with the need for immunosuppressive therapy"

**Suppl. Material:**

**Quantification of polyreactive immunoglobulin G**

Patients’ serum samples from Hannover Medical School were cryo-conserved at below -20°C. Serum samples from external centers were cryo-conserved according to local protocols and sent frozen to Hannover Medical School for centralized quantification of pIgG. Samples were pseudonymized for autoantibody-testing and observers were blinded to any clinical information.

Quantification of pIgG using an ELISA to quantify reactivity to a peptide and BSA as blocking agent was performed as published^1^. In short, 0.01 µg of HIP1R fragment per well was bound to 96 well ELISA plates over night at 4 °C. Plates were blocked with TBS and 5 % BSA for 30 minutes. Plates were washed with TBS with Tween20® 0.05 % (TBST) once. Serum samples were diluted 1:101 (v/v) in TBS and 5 % BSA and 100 µl per well were added to the ELISA plate and incubated for two hours. Plates were washed three times with TBST and incubated with a secondary rabbit anti-human anti-IgG antibody labeled with horseradish peroxidase for 30 minutes. Three washing steps with TBST were performed and 3, 3', 5, 5' tetramethyl benzidine (BioLegend, San Diego California) was added for 30 minutes for color reaction. Reaction was stopped with sulfuric acid. Optical density was read at 450 nm using an ELISA reader (Tecan Sunrise-Basic, Grödig, Austria). Sera of five patients with gradual increase in pIgG reactivity were measured in every experiment and used to compute a standard curve. Arbitrary units (AU) were calculated from the equation of the standard curve. Measurements were performed in Hannover, Germany.

As different AU dependent on center and storage duration were demonstrated, a normalization for these factors (referred to as normalized AU (nAU)) was performed as published^1^.

**Immunofluorescence testing**

IFT was performed by experienced technicians using the recommended methodology of the guidelines issued in 2004 by the Committee for Autoimmune Serology of the International Autoimmune Hepatitis Group^2^. Samples were pseudonymized for autoantibody-testing and observers were blinded to any clinical information. ANA, anti-SMA, anti-LKM and anti-LC1 were detected by IIF on sections of frozen rodent liver, stomach and kidney sections. Briefly, a commercial rodent multi-organ substrate panel (kidney, liver and stomach) was used (LKS Rat wrapped Standard Kit, Aesku.Diagnostics GmbG & Co. Wendelsheim, Germany). The sera were diluted, starting with a dilution of 1:20 up to 1:320, and applied to the slide to cover the entire tissue section and allow binding of the autoantibodies to the substrates. After washing, the sample was exposed to a second fluorochrome-labeled antibody. Finally, once washed again, the slides were examined under fluorescence microscope (Olympus BX60 Microscope, Evident Europe GmbH, Germany), and the antibody staining pattern was evaluated and interpreted accordingly to the guidelines^2^.
